# The Cognitive Safety Net: Comparing Human and AI Diagnostic Reasoning during Complex Clinical Situations

**DOI:** 10.1101/2025.10.06.25335641

**Authors:** Antonin Audion, Mathieu Henkeme, Baptiste Balanca, Marc Lilot, Thomas Rimmelé, Ismail Abaakil, Jean-Christophe Cejka

## Abstract

**Background:** **Diagnostic** error in high-stakes clinical environments remains a significant cause of preventable harm. While a new generation of customisable digital cognitive aids (cDCAs) has shown a capacity to improve performance, achieve robust competence, and double learning retention, the potential for artificial intelligence (AI) to augment the foundational, anticipatory reasoning that precedes action is not well understood. This study aims to compare the diagnostic reasoning strategies of experienced anaesthesiology residents with those of a large language model (LLM) during a simulated, complex and realistic anaesthesiology scenario.

**Methods:** We conducted a comparative analysis within a high-fidelity simulation randomised controlled trial (Anticipamax, NCT06487208). Thirty-four experienced anaesthesiology residents and a conversational LLM (ChatGPT-4) managed a perioperative shock of deliberately multifactorial aetiology. Diagnostic lotteries—sets of hypotheses with assigned plausibility scores—were collected before and after the simulation. We implemented a novel analytical framework based on the social choice **Condorcet method**, to rank not only individual hypotheses but also to compare the complete diagnostic strategies as the case evolved.

**Results:** The AI and residents demonstrated distinct reasoning profiles. Initially, the AI produced an exhaustive, non-hierarchical analysis, correctly identifying septic shock among its top, similarly-scored hypotheses. Residents, in contrast, employed a pragmatic, focused strategy, prioritising immediate surgical risks and unanimously identifying an experience-based risk (gas embolism) that the AI systematically overlooked, and consistently reserved a portion of their reasoning for uncertainty, termed ‘Place for Doubt’. After the clinical evolution, both converged on septic shock. A ‘complex scrutiny’ analysis of the overall strategies revealed that the **residents’ focused and adaptive reasoning was consistently ranked as strategically superior** to the AI’s exhaustive but diluted approach.

**Conclusions:** AI demonstrates a powerful capacity for broad diagnostic anticipation, acting as a potential **safeguard against premature diagnostic closure**. Experienced residents exhibit a strategically superior reasoning process in its focus and adaptation. Our findings support a **powerful synergy** where the AI serves as a ‘Cognitive Safety Net’ to augment, not replace, the contextualised judgment of the human practitioner.

**Research in Context:** *What is already known on this topic:*

- Human error in healthcare is a global prominent cause of death.
- **‘Traditional’ cognitive support tools** (e.g., paper checklists) have been shown to improve technical skills during medical crises, but their impact on non-technical skills is limited and their clinical adoption remains low.
- A new generation of customisable digital cognitive aids (**cDCAs**) can significantly improve both technical and non-technical performance, fostering better team management and crisis resolution.
- Information on how clinicians deliver the best **anticipatory clinical reasoning** is scarce.
- Recent work comparing machine-learning models to clinicians in trauma triage found comparable accuracy but only moderate agreement, suggesting a collaborative paradigm and motivating deeper analyses of the reasoning process itself.
- However, a **critical gap remains in understanding the underlying nature of the diagnostic reasoning strategies** that lead to these outcomes. The ‘how’ of human and AI reasoning, especially in dynamic, anticipatory clinical tasks, is not well understood.

*What this study adds:*

- This is the first study to directly compare in action the diagnostic reasoning strategies o*f clinicians and a large language model* (AI).
- It introduces a novel analytical framework based on the **Condorcet** social choice method to **move beyond simple performance scores** and rigorously **model and rank the overall quality of diagnostic strategies** in a simulated daily complex situation.
- The findings support a model of **human-AI complementarity**, where the AI excels at broad, exhaustive analysis, while clinicians demonstrate a superior, focused, and adaptive strategic reasoning, suggesting the human’s role as a **meta-cognitive supervisor** of AI-driven exhaustive but ‘diluted’ insights.

## Introduction

Medical error in complex, high-pressure environments like the operating theatre remains a leading cause of preventable harm and mortality^1,2^. Many such errors stem not from a fundamental lack of knowledge, but from failures in cognition under duress, where cognitive overload can lead to diagnostic fixation and critical omissions. Inspired by high-reliability industries, healthcare has adopted cognitive support tools to mitigate these risks.

The first generation of these tools, primarily paper-based checklists and static algorithms, proved effective at improving adherence to technical protocols but showed limited impact on crucial non-technical skills such as team communication and dynamic decision-making^3^. More recently, customisable digital cognitive aids (cDCAs) like MAX have overcome many of these limitations, with multiple randomised controlled trials demonstrating their capacity to double learning retention^4^, significantly enhance both technical *and* non-technical performance during simulated high-stakes scenarios^5–9^, and cut total error—defined as the sum of systematic deviation from standards (bias^2^) and inter-individual variability (variance)—by 75%^10^.

This progress raises a new scientific question: now that we have tools to better execute tasks, can we augment the very foundation of clinical action—the anticipatory reasoning process itself? The emergence of powerful artificial intelligence (AI) offers a potential pathway. Recent prospective evidence in pre-hospital trauma care showed that a machine-learning model and clinicians achieved similar predictive performance but only moderate agreement, with complementary error profiles; crucially, combining human and algorithm yielded a net sensitivity gain^11^. This strengthens the rationale for our strategy-level comparison. Rather than testing ‘who is more accurate’, our study models ‘how each system reasons’ under uncertainty, using a unique analytical framework derived from social choice theory—the Condorcet method^12^. By **comparing not just static lists, but** the evolution of ranked priorities and **the overall strategic quality of diagnostic ‘lotteries’**, we can directly compare the judgments of clinicians and of a generalist LLM (ChatGPT-4).

Dissecting the strengths, weaknesses, and potential synergies of these two forms of intelligence is a critical step towards a future where clinical reasoning is not replaced, but robustly augmented.

## Methods

### Context

This comparative analysis was conducted as an ancillary study using data from the ‘Anticipamax’ randomised controlled trial (ClinicalTrials.gov ID NCT06487208)^13^. The parent trial’s protocol included the systematic collection of diagnostic lotteries from participants. The main trial included 34 residents, and the present human-AI comparative analysis focused on the diagnostic lotteries collected from the first 25 consecutively enrolled participants, a sample size defined by the logistical constraints of the parallel master’s degree project by AA. The standardised, high-fidelity simulation scenario is detailed in ‘Supplementary Appendix 1’. It involved the management of a poly-pathological patient who subsequently developed a state of shock intraoperatively. The aetiology of the shock was deliberately designed to be multifactorial, with no single ‘correct’ answer revealed at the end of the scenario.

### Ethics statement

The Anticipamax study used high-fidelity simulation with volunteer anaesthesiology residents. No patient data were used. Participants provided informed consent to participate and to the anonymised use of their responses for research. According to local regulation, the project was accepted by the Ethical Committee of the French Society of Anaesthesiology-Intensive Care (CERAR, *‘Comité d’Éthique pour la Recherche en Anesthésie-Réanimation’*) as educational research without patient involvement (IRB 00010254- 2024– 019).

### Study Design

Participants were briefed on the patient’s clinical history as part of a pre-interventional handover from a departing ‘colleague’ (same simulation instructor all along the trial). Immediately following this briefing, they were instructed to compile a list of foreseeable complications, assigning a plausibility score to each. This process was repeated upon completion of the simulated scenario, yielding two diagnostic lotteries—lists of diagnoses, each assigned an estimated plausibility—for each participant, designated ‘Before’ and ‘After’.

### Participant Groups

The study included two participant groups. The human cohort comprised 34 experienced anaesthesiology residents aged 26 to 30 (second to fifth year of medical specialty training in the French system (DES 2–5), roughly corresponding to specialty training years ST3–ST6/7 in the UK system).This specific population was chosen deliberately. Being in an advanced stage of their curriculum, they possess a comprehensive and, critically, an up-to-date knowledge of current guidelines and pathophysiology. This minimises the risk of knowledge deficit or simple forgetting being a confounding factor, allowing the study to focus specifically on the structure and strategy of their reasoning process when faced with uncertainty. The AI counterpart was the ChatGPT-4 large language model (OpenAI, California, USA). For the ‘Before’ analysis, it was prompted with the identical text-based case information provided to the residents. For the ‘After’ analysis, to ensure a fair comparison, the key chronological events of each unique human simulation session were transcribed and used to prompt the AI for its revised diagnostic lottery.

### Analytical Framework: The Condorcet Method

To move beyond simple performance scores, we developed a novel framework based on the Condorcet method, a robust tool from social choice theory^12^. This approach allowed us to model and rank not just individual hypotheses but entire diagnostic strategies. Each participant’s list of ranked diagnoses was treated as a ‘diagnostic lottery’—a portfolio of plausibilities reflecting their reasoning under uncertainty.

The Condorcet method was deliberately chosen as it allows for the evaluation of holistic diagnostic strategies rather than isolated variables. By analyzing pairwise preferences within each diagnostic lottery, this method preserves the relational structure of clinical reasoning, enabling a comparison of the overall strategic quality and adaptability of human versus AI judgment under uncertainty

Following **data standardisation**, where raw diagnostic terms were algorithmically harmonised using a predefined lexicon, our analysis proceeded in two stages.

First, a ‘**Simple Scrutiny’** ranked individual hypotheses by conducting pairwise ‘duels’ to identify which diagnoses were most consistently preferred across all lotteries.

Second, a ‘**Complex Scrutiny’** assessed the overall quality of each complete diagnostic strategy (lottery) by comparing it against every other strategy, yielding a robust ranking of the most strategically sound reasoning approaches.

This dual-level analysis reveals the structure, focus, and adaptability of reasoning for both clinicians and the AI. The detailed computational protocol, theoretical justification, and mathematical underpinnings of this method are provided in the Supplementary Appendix 2 and 3.

## Results

### Ranking of Diagnostic Hypotheses

The Simple Condorcet Scrutiny revealed a clear evolution in diagnostic reasoning. Before the case management, clinicians ranked **haemorrhagic shock** highest, whilst the AI prioritised **septic shock**. After the crisis, both groups converged on **septic shock** as the dominant hypothesis.

Tables 1.1 and 1.2 show the top four most dominant hypotheses for each group before (post-briefing) and at the very end of the simulation.

**Table 1.1:** Ranking of Diagnostic Hypotheses (Simple Condorcet Scrutiny) before the simulation.

| Before the Simulation |  |  |  |  |
| --- | --- | --- | --- | --- |
| Rank | Residents - Before | Score | AI - Before | Score |
| 1 | Haemorrhagic Shock | 48 | Septic Shock | 54 |
| 2 | Septic Shock | 38 | Acute Hepatic Failure | 52 |
| 3 | Gas Embolism | 22 | Haemorrhagic Shock | 48 |
| 4 | Place for Doubt | 5 | Respiratory Decompensation | 44 |

**Table 1.2:**
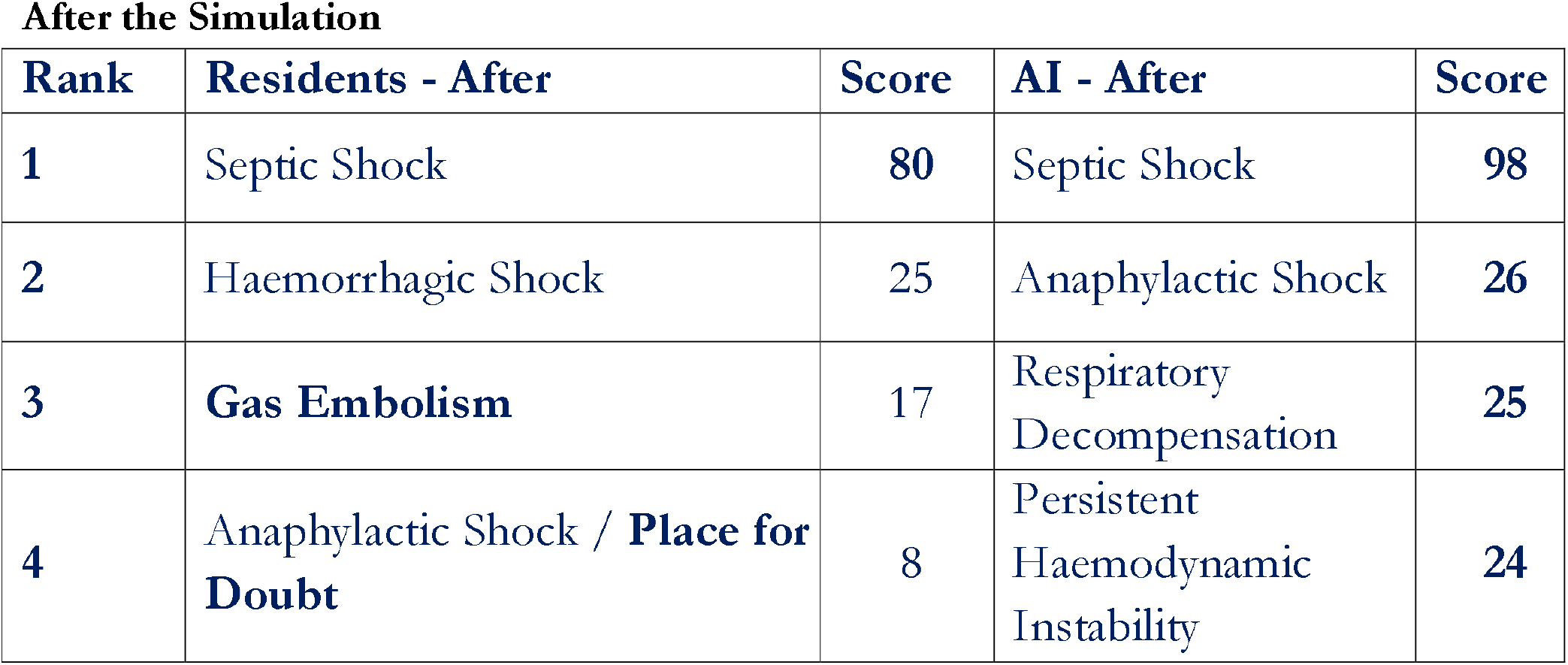
Ranking of Diagnostic Hypotheses (Simple Condorcet Scrutiny) after the simulation.

| After the Simulation |  |  |  |  |
| --- | --- | --- | --- | --- |
| Rank | Residents - After | Score | AI - After | Score |
| 1 | Septic Shock | 80 | Septic Shock | 98 |
| 2 | Haemorrhagic Shock | 25 | Anaphylactic Shock | 26 |
| 3 | Gas Embolism | 17 | Respiratory<br>Decompensation | 25 |
| 4 | Anaphylactic Shock / **Place for Doubt** | 8 | Persistent<br>Haemodynamic<br>Instability | 24 |

**Table 2:**
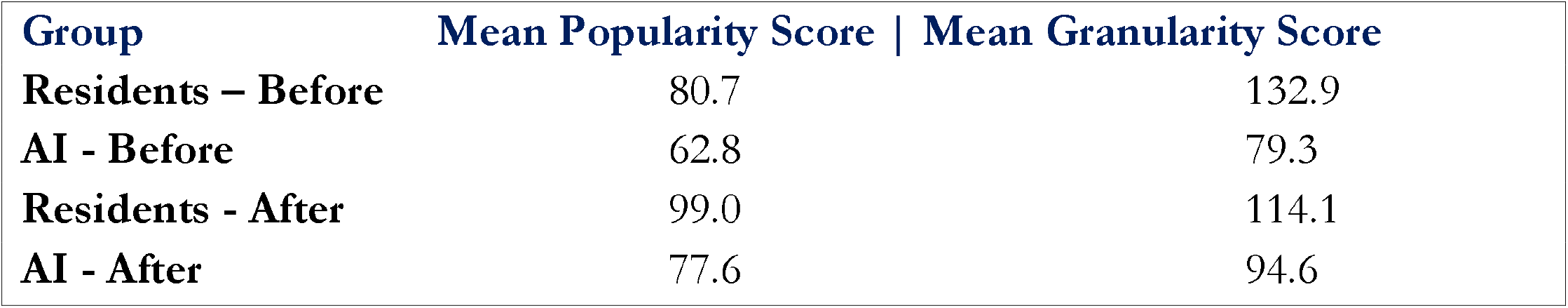
Mean Strategic Performance Scores (POPU & GRANU)

### Ranking of Overall Diagnostic Strategies

The Complex Scrutiny, evaluating each entire diagnostic lottery, showed that the clinicians’ strategies were, on average, ranked higher than the AI’s.

The clinicians demonstrated a marked improvement in their strategic popularity score after the crisis (80.7 to 99.0), indicating a **highly effective adaptation** to the clinical data.

### Visualisation of Diagnostic Reasoning Structures - Chord Graphic representation

To visualise the underlying structure and dynamics of the diagnostic reasoning, the pairwise dominance relationships from the Simple Condorcet Scrutiny were rendered as Chord diagrams (Figure 1). These diagrams function as maps of the collective diagnostic mind of each group. In these maps, a **dominant hypothesis** appears as a node with numerous, thick *outgoing* arrows, indicating it consistently won its pairwise duels against other hypotheses. A **highly contested hypothesis**, in contrast, shows both significant incoming and outgoing arrows, marking it as a central but debated topic in the reasoning process. Finally, the **overall structure** reveals the nature of the group’s reasoning: a simple, hierarchical diagram with one clear dominant node suggests strong consensus and diagnostic convergence, whereas a complex, web-like diagram indicates a broader and less hierarchical differential.

**Figure 1:**
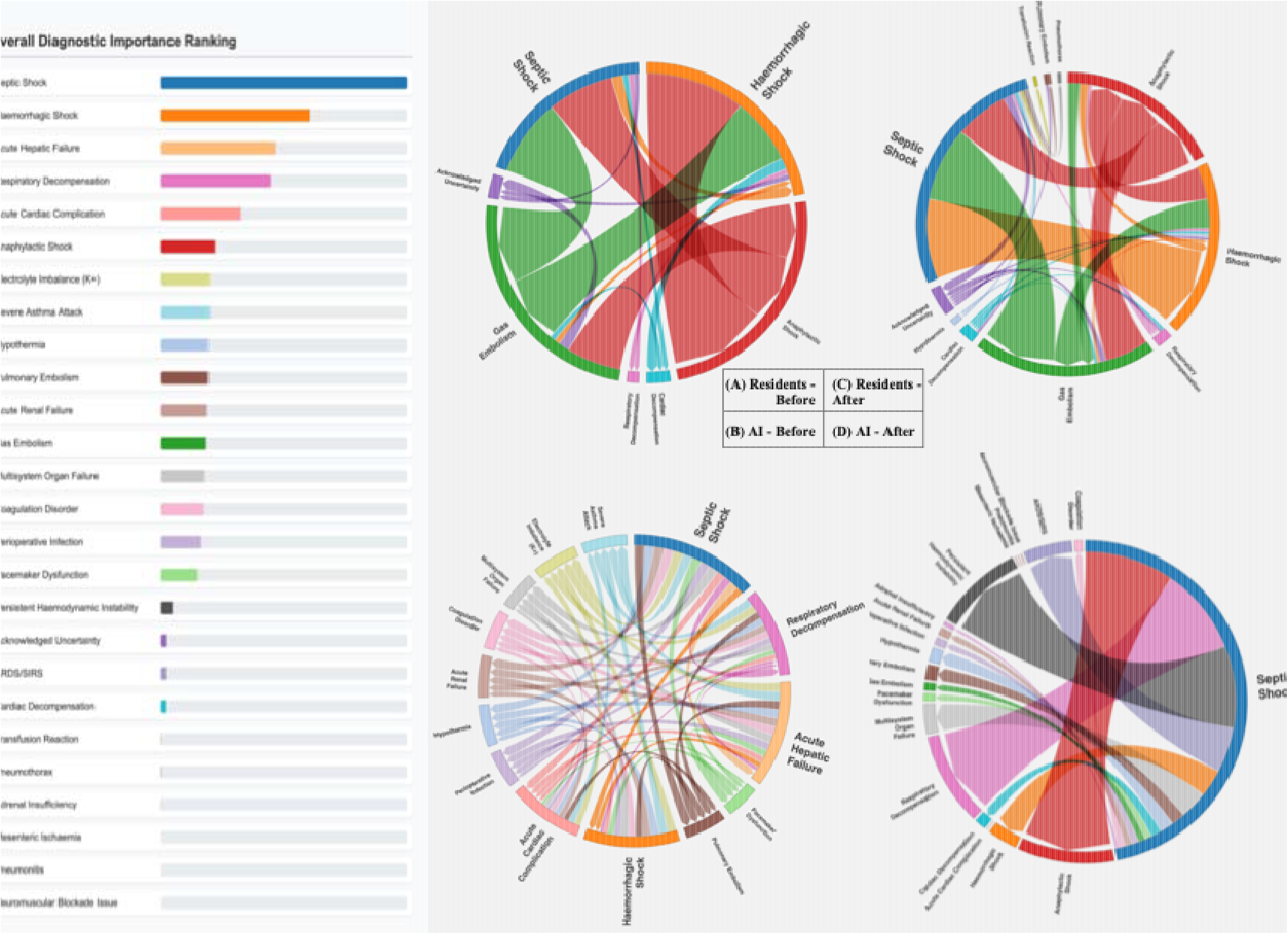
Comparative Analysis of Diagnostic Reasoning Structures via Condorcet Scrutiny. The figure presents a 2x2 grid of Chord diagrams, visualising the results of the Simple Condorcet Scrutiny for the four experimental groups. Each main arc on the circumference represents a single diagnostic hypothesis, with colours kept consistent across all four diagrams for direct comparison. The directed arrows between arcs illustrate the dominance relationship from a source (dominant) to a target (dominated) hypothesis. The thickness of each arrow is proportional to the total number of pairwise ‘duels’ won by the source hypothesis against the target hypothesis across all diagnostic lotteries within that group. The panels show:

- **(A) Experienced Residents - Before:** The baseline reasoning structure of the anaesthesiology residents prior to the simulated event.
- **(B) AI - Before:** The baseline reasoning structure of the AI based on the same initial case information.
- **(C) Experienced Residents - After:** The adapted reasoning structure of the residents following the management of the simulated event.
- **(D) AI - After:** The adapted reasoning structure of the AI after being prompted with the evolving clinical data.

## Discussion

The fundamental nature of clinical reasoning is not a simple dichotomy between right and wrong, but a dynamic interplay of strategy and uncertainty^14,15^. This study reveals that human and artificial intelligences do not simply compete on accuracy but operate as distinct, **complementary reasoning engines**. This discovery moves the conversation **beyond a simplistic human-versus-machine narrative** to a more sophisticated understanding of human-AI collaboration in medicine, where the goal is not replacement, but the creation of a more robust, resilient form of augmented clinical intelligence.

### Unveiling Two Minds at Work: The Pragmatic Filter vs. the Exhaustive Map

The core of our study lies in the dissection of two fundamentally different approaches to resolving diagnostic uncertainty.

### The AI’s approach is one of exhaustive, high-fidelity transmission

Its initial diagnostic lottery, which assigned nearly equal plausibility scores to its top hypotheses (septic shock at 54, acute hepatic failure at 52, haemorrhagic shock at 48 and respiratory decompensation at 44), exemplifies a systematic, probabilistic mapping of the data. This lack of a strong initial hierarchy, while strategically less decisive, provides a powerful breadth reducing the initial ‘universal’ (i.e. all existing diagnoses) uncertainty to a manageable set of possibilities, thereby, acting as a robust safeguard against cognitive fixation and premature diagnostic closure.

In stark contrast, the residents’ reasoning is a process of **aggressive, context-sensitive filtering**. Their lotteries were sharply hierarchical, with haemorrhagic shock (48) clearly dominating septic shock (38). Crucially, this focused strategy included the unanimous identification of gas embolism (22)—a context-specific risk entirely absent from the AI’s initial analysis. This finding exemplifies a classical *Naturalistic Decision Making* (NDM) model^15^, where experts recognize familiar patterns associated with the specific context-liver surgery, rather than analytically deducing solutions. This intuitive pattern-matching, born from embodied experience, represents a first, distinct advantage of human cognition.

The most profound distinction, however, resides in the **fourth diagnostic choice**. While the AI proposes another specific clinical entity with a similar plausibility (Respiratory Decompensation, score 44), the residents formalize a core principle of expert cognition: the **‘Place for Doubt’** (score 5). This is not a failure to hypothesize, but rather an efficient **metacognitive strategy**—the explicit allocation of cognitive resources to residual uncertainty. After the crisis, even with a near-certain primary diagnosis of septic shock, the residents maintained this ‘Place for Doubt’ in their final lottery (tied for 4th rank, score 8), demonstrating that this cognitive safety net persists even as certainty increases.

This concept of complementary reasoning profiles is further substantiated by recent work in trauma care^11,16^, which found only moderate agreement and distinct error patterns between machine learning models and clinicians, with combined performance exceeding that of either alone.

### An Information-Theoretic Perspective: Managing Clinical Entropy

These contrasting strategies can be powerfully framed using the lens of information theory^17^. A complex clinical situation represents a system of high entropy—a measure of disorder and uncertainty. Effective clinical reasoning **is a battle to reduce this entropy** and find the ‘signals’ (i.e. the correct diagnoses) within a sea of noise^18^. Humans and AI employ radically different, yet complementary, strategies to win this battle. The AI’s multi-hypothesis lottery represents a state of relatively high entropy, acting as a ‘**broadband communication channel’** that faithfully transmits all potential signals. Conversely, the residents’ focused lottery represents a low-entropy state; the human expert acts as a ‘**highly selective adaptive filter’**, using the *non-linearities of experience* to amplify the perceived signal and suppress the noise, thus making a decision actionable. Interestingly, the ‘Place for Doubt’ is the human’s unique method for managing **irreducible entropy**—acknowledging the noise that always remains in a complex biological system.

The **Chord diagrams** offer a **phenomenological visualization of these entropic states**. The complex, web-like structure of the AI’s ‘Before’ diagram (Fig. 1B), with its numerous, fine, balanced arrows, is the **graphical signature of a high-entropy**, non-hierarchical state. Conversely, the simpler, more radial structure of the residents’ ‘Before’ diagram (Fig. 1A), dominated by a few thick arrows, visually represents a **low-entropy state** of strong consensus and clear **strategic focus**.

### The Synthesis: A New Model for Augmented Reasoning —The Cognitive Safety Net

Recognizing these distinct strategies allows us to propose a new model of collaboration: the **‘Cognitive Safety Net’**. In this model, the true value of AI is not to provide a ‘better’ answer, but to serve as an exhaustive knowledge base that prevents the primary failure mode of expert cognition: fixation bias. Its broad analysis acts as the cognitive safety net, prompting the clinician to consider possibilities outside their immediate focus.

The human practitioner, however, remains the ultimate strategist and **meta-cognitive supervisor**. They are responsible for applying ‘contextual wisdom’ from experience-based heuristics—context, experience, and an understanding of the patient’s trajectory— to the probabilistic outputs of the AI and managing uncertainty—a process exemplified by their ‘Place for Doubt’. The clinician’s role evolves **from** being a **repository of knowledge to** being an **expert arbiter of it**. This supervisory capacity is critical, as a paradox of AI in medicine is that its very logical coherence can induce anchoring bias in the user. The clinician’s role is therefore to step back and maintain a global perspective.

This model also clarifies the **distinction between knowledge and competence**. The AI provides structured information (‘knowledge’), but it cannot manage a team, perform a procedure, or handle the immense cognitive load of a real high-stakes situation (‘competence’). As Kassirer articulated, true clinical competence is not merely the possession of knowledge, but its effective application under conditions of uncertainty^14^—a process that remains fundamentally human. This transition from knowledge to competence is precisely where the clinician is most vulnerable to cognitive overload, a challenge that makes the supervisory role over an AI untenable without cognitive support.

This is where efficient procedural cognitive aids become essential. In an optimal workflow, the AI assists in the diagnostic phase (‘*What is happening’*), after which a proven, action-oriented cDCA guides the team through validated procedures (‘*How to manage it*’). By offloading the procedural burden, these tools free the clinician’s cognitive bandwidth to effectively supervise all necessary data—which include the AI’s insights— and manage the evolving clinical situation. Current evidence already indicates that tailored cDCAs can considerably decrease human errors, improve both technical and non-technical skills across a wide range of situations^3,5–9,13^, and double learning memorization^4^. Thus, further enhancing human capacity with this dual-support system—AI for enriching reasoning, cDCAs for contextualized action—is a pragmatic, ethical, and highly efficient path to reducing error in healthcare.

### Implications and Future Horizons: Towards a Risk-Weighted Analysis

The ‘Cognitive Safety Net’ model suggests that AI integration should prioritize tools that broaden the diagnostic horizon and highlight not just the most likely events, but also the most critical ‘black swan’ scenarios (*‘What if’)*—those that are highly unlikely but would be catastrophic. This would elevate the AI from an analytical tool to a true partner in navigating high-stakes uncertainty. In practice, this synergy could be channelled through cDCAs, presenting context-specific workflows while embedding AI-derived insights among other contextualized data, in a usable form at the bedside.

### Strengths and Limitations

The primary strength of this study is its novel methodology. By using a realistic scenario with deliberate ambiguity and applying the **Condorcet method** to ‘diagnostic lotteries’, we shifted the analytical focus **from the diagnostic result to the reasoning process**. Our multi-layered evaluation—ranking individual hypotheses via ‘Simple Scrutiny’ and entire strategies via ‘Complex Scrutiny’—provided a granular framework for modeling the *how* of clinical reasoning, not just the *what*.

A potential limitation is the use of a generalist LLM (ChatGPT-4). However, this was a deliberate choice to compare the specialized, experience-driven reasoning of clinicians against a powerful, non-specialist form of general logical inference, revealing their foundational differences. Future research should compare these findings with medically fine-tuned models.

Similarly, we chose experienced residents to focus on the structure of reasoning in a cohort with a highly current knowledge base, neutralizing the variable of knowledge obsolescence. Our research widely confirms that when equipped with our cDCAs, this population’s performance is considerably enhanced^5–10,13^, mitigating a potential performance gap with senior clinicians, who remain subject to human error in action.

Another potential limitation is the sample size discrepancy between the full Anticipamax trial cohort (N=34) and the cohort included in this specific human-AI reasoning analysis (N=25). This difference is due to the nature of our work as a planned ancillary study, with a timeline constrained by a parallel master’s degree project by AA. However, we argue that this does not invalidate our conclusions for two reasons. First, a sample of 25 experienced residents is substantial for a study focused on modeling the process and structure of reasoning, rather than achieving statistical power for a clinical outcome. Second, the reasoning profiles we identified—the AI’s exhaustive mapping versus the residents’ pragmatic filtering—emerged with remarkable consistency and clarity within this cohort. It is therefore unlikely that the inclusion of the final nine participants would have fundamentally altered these distinct strategic signatures.

## Conclusion

This work reveals that **AI is not poised to replace the clinical mind**, but **to augment it**. While the focused, adaptive strategy of clinicians remains superior in a dynamic, high-stakes situation, AI provides a powerful, broad-based anticipatory analysis that complements human reasoning.

By integrating AI as a ‘Cognitive Safety Net’ within a human-centred workflow, we can leverage the strengths of both intelligences to create a more robust and safer system of care. Ensuring future generations of AI are designed with this synergistic, human-in-the-loop principle is not only the best choice for quality of care, but also a profound ethical imperative.

## Data Availability

All data produced in the present study are available upon reasonable request to the authors

## Supplementary Information accompanies this paper

- **Supplementary Appendix 1**. Case Scenario Briefing.
- **Supplementary Appendix 2**. Methodological Framework and Rationale.
- **Supplementary Appendix 3**. Detailed Analytical Protocol – Condorcet scores.

## Supplementary Appendix 1: Case Scenario Briefing

**Patient:** Mrs. M., 51-year-old female.

**Procedure:** Right hepatectomy via laparotomy.

**Clinical Background:** You are the anaesthetist taking over the care of Mrs. M.

The patient was recently diagnosed with colonic adenocarcinoma with two synchronous hepatic metastases. Given her age and otherwise limited comorbidities, the multidisciplinary team (MDT) meeting approved a surgical approach with curative intent.

Consequently, one month ago, she underwent a right colectomy plus a radiofrequency ablation (RFA) of the hepatic dome lesion. The postoperative course was complicated by haemorrhagic shock from the RFA site, requiring embolization of the right hepatic artery. This subsequently led to purulent necrosis of the right lobe of the liver.

Following the failure of both radiological drainage and empirical antibiotic therapy (cefotaxime and metronidazole), and in the context of persistent, low-grade sepsis, an indication for today’s right hepatectomy was confirmed.

### Past Medical History

- **Medical**
  - Syncope secondary to second-degree atrioventricular block (AVBII) in 2022, leading to the insertion of a pacemaker (last checked in 2023).
  - Familial osteoporosis.
  - Post-smoking chronic obstructive pulmonary disease (COPD), with significant winter wheezing.
- **Surgical**
  - Appendicular peritonitis at age 38.
  - Femoral fracture from a fall (secondary to syncope) in 2022.
  - Received a blood transfusion in 2022.
- **Social & Allergies:**
  - 30-pack-year smoking history; quit 2 years ago following the diagnosis of her heart block.
  - No significant alcohol intake.
  - No known drug allergies (NKDA).

## Supplementary Appendix 2: Methodological Framework and Rationale

### 1. The Rationale for a Condorcet-Based Framework: A Practical Example

To illustrate the robustness of the ‘Condorcet method’ compared to ‘simpler voting systems’, consider this clinical example.

After a patient evaluation, three main plausible diagnoses arise (say A, B and C), and five clinicians are asked to provide their own ‘diagnostic lotteries’ (Table A1):

**Table A1:** Example Diagnostic Lotteries from Five Clinicians.

| Clinician | Plausibility for A | Plausibility for B | Plausibility for C | First Choice |
| --- | --- | --- | --- | --- |
| Clinician 1 (L1) | 60 | 30 | 10 | A |
| Clinician 2 (L2) | 60 | 30 | 10 | A |
| Clinician 3 (L3) | 10 | 60 | 30 | B |
| Clinician 4 (L4) | 30 | 10 | 60 | C |
| Clinician 5 (L5) | 30 | 10 | 60 | C |
| **Total Plausibility:** | **190** | **140** | **170** |  |

#### The Failure of Simple Metrics

A simple analysis based on **total plausibility score** would declare **Diagnosis A** the winner (Total Score: 190). Similarly, a **plurality vote** based on first preferences results in a tie between Diagnosis A and C (2 votes each), failing to identify a clear winner and ignoring the nuances of the other preferences.

Both results are heavily skewed by the strong conviction of a minority (Clinicians 1 & 2).

#### The Simple Condorcet Scrutiny: A Cyclical Paradox

When we analyse the pairwise ‘duels’ based on how many clinicians prefer one option over another, a paradox emerges:

- **Duel A vs. B**: Clinicians 1, 2, 4, and 5 prefer A to B. Clinician 3 prefers B to A. - > **A wins 4-1**.
- **Duel B vs. C**: Clinicians 1, 2, and 3 prefer B to C. Clinicians 4 and 5 prefer C to B. -> **B wins 3-2**.
- **Duel C vs. A**: Clinicians 3, 4, and 5 prefer C to A. Clinicians 1 and 2 prefer A to C. -> **C wins 3-2**.

This creates a cycle of preferences (**A > B > C > A**), known as a **Condorcet Paradox**. There is no single winning diagnosis that can defeat all others, leaving the result undecided.

#### The Complex Scrutiny: Identifying the Winning Strategy

This is where the power of comparing entire lotteries becomes evident. When we run the Complex Scrutiny (POPU and GRANU scores) on the five lotteries:

**Table A2:** Complex Scrutiny Results for the Example Lotteries.

| **Lottery (Clinician)** | **Popular Score (POPU)** | **Granular Score (GRANU)** | **Rank** |
| --- | --- | --- | --- |
| **Clinicians 1&2 (L1, L2)** | **4** | **6** | **1** |
| Clinician 3(L3) | 2 | 4 | 3 |
| Clinicians 4&5 (L4, L5) | 2 | 3 | 4 |

The winning strategy is clearly that of **Clinicians 1 and 2**. Their lottery—{**A: 60, B: 30, C: 10**}—is identified as the most robust because it represents the best strategic compromise: it shows strong conviction for the most likely diagnosis (A) while still assigning a reasonable plausibility to a key differential (B), making it the most resilient option in pairwise comparisons against more polarised strategies. This example demonstrates how our method moves beyond simple votes to identify the most coherent and strategically sound reasoning process.

### 2. Theoretical Foundation: Randomized Condorcet Voting and the Lottery Space

The analytical framework of this study is grounded in the principles of **Randomized Condorcet Voting**, a mathematically elegant solution to the paradoxes inherent in collective decision-making.

The fundamental problem of standard voting systems, including the simple Condorcet method, is the potential for **intransitivity of collective preferences**. As shown in the example above, it is possible for a group to prefer A over B, B over C, yet prefer C over A, resulting in a cycle with no clear winner. This paradox has been a central challenge in social choice theory for centuries.

The theoretical innovation, formalised in recent mathematical literature, is to resolve this paradox by changing the ‘solution space’. Instead of seeking a winner from the set of discrete *candidates* (our diagnoses), the method seeks a winner from the infinite set of *lotteries*. A lottery is a probability distribution over the set of candidates. In our study, each participant’s diagnostic list, with its assigned plausibilities, is a lottery.

To apply the Condorcet principle to this new space, a method for comparing two lotteries, P and Q, is required. The preference of the electorate for P over Q is determined by the probability that a candidate randomly drawn from lottery P is preferred by a majority to a candidate randomly drawn from lottery Q. This elevates the comparison from the level of single options to the level of overall strategies.

The central theorem underpinning this method is profound: under general conditions, there is **always a unique Condorcet-winning lottery**, even when a winning candidate does not exist. The paradox vanishes. By moving to a probabilistic solution space, the existence and uniqueness of a ‘best choice’ are guaranteed.

This is the justification for our two-level analysis. The **Simple Scrutiny** seeks the winning diagnosis but may fail due to paradoxes. The **Complex Scrutiny** operates on the higher level of lotteries, providing a more stable and robust assessment that identifies the superior overall reasoning strategy.

### 3. Connecting Theory to Clinical Practice: A Bayesian and Decision-Theoretic Framework

The application of this mathematical framework^19^ to clinical reasoning^20^ is justified by two core principles:

#### 3.1 The Bayesian Clinician: Clinical reasoning is an inherently Bayesian process

A clinician does not operate in a world of certainties, but of probabilities. The plausibility scores assigned in a ‘diagnostic lottery’ can be seen as subjective probabilities—Bayesian priors—that are continuously updated as new information becomes available (a change in vital signs, a lab result). A clinician’s expertise lies in their ability to effectively update these probabilities in real time. Our plausibility-weighted Condorcet method is therefore a formal means of aggregating these evolving, probabilistic beliefs to find a robust group consensus.

#### 3.2. Decision-Making as a Bet

As decision scientist and former professional poker player Annie Duke argues^21^, expert decision-making under uncertainty is not about finding the ‘*truth*’, but about making the ‘**best possible bet**’ given incomplete information.

- Seeking a single winning diagnosis is akin to going ‘*all-in*’ on a single hand. It is a brittle strategy that ignores the spectrum of possibilities.
- Reasoning in ‘lotteries,’ by contrast, is analogous to managing a portfolio of possibilities. The clinician is not betting on one outcome but is distributing their ‘*capital of belief*” across a range of potential futures.

Our Complex Scrutiny, therefore, does not reward the best guess; it rewards the best **betting strategy**. It identifies the reasoning process that is the most robust, balanced, and likely to succeed across the full range of potential outcomes. This aligns perfectly with the reality of high-stakes clinical work, where the goal is not to be right once, but to be consistently resilient in the face of uncertainty.

## Supplementary Appendix 3: Detailed Analytical Protocol – Condorcet scores

This document details the step-by-step computational protocol used to analyse the diagnostic lotteries and generate the Condorcet-based rankings.

**Condorcet scores** were originally designed by Nicolas de Condorcet (1743-1794, scientist, mathematician, philosopher, politician) to identify the candidate who best reflects the collective preference of an electorate, i.e. the one who would defeat every other rival in a head-to-head contest^12^.

### 1. Data Standardisation

- The initial step consisted of standardising all diagnostic terms to ensure semantic consistency.
- Raw diagnostic lotteries from all participants (human and AI) were compiled.
- Each diagnostic term was compared against a master reference file of approved standardised terms.
- If a term did not have an exact match, it was compared against a second file containing known variants. If a match was found, the term was replaced by its corresponding standardised term.
- Any remaining non-standardised terms were flagged for manual review and categorisation by an experienced clinician (JCC).
- A final verification of the standardisation for each term was automatically performed by a purpose-configured GPT (ChatGPT4, OpenAI, California).

### 2. Simple Condorcet Scrutiny: Hypothesis Ranking

This analysis aimed to rank individual diagnostic hypotheses based on their dominance.

- **Creation of the Normalised Plausibility Matrix ([PN])**: A matrix was constructed where each row *i* represents a participant’s lottery and each column *j* represents a standardised hypothesis. The cell *PN(i,j)* contains the plausibility score for that hypothesis in that lottery (or 0 if absent).
- **Creation of the Pairwise Wins Matrix ([ARROWS])**: An *N x N* matrix (where *N* is the number of unique hypotheses) was created. For each lottery, a ‘duel’ was conducted between every pair of hypotheses *(j, k)*. If *PN(i,j) > PN(i,k)*, the cell *ARROWS(j,k)* was incremented by 1. The final matrix thus contains the total number of times each hypothesis ‘won’ a direct comparison against another in terms of quoted plausibility. In the spirit of the Condorcet score, this is equivalent to counting the number of ‘arrows’ originating from a *‘dominant’* hypothesis and pointing towards *‘dominated’* hypotheses.
- **Calculation of Final Hypothesis Ranking**: The total Condorcet score for each hypothesis *j* was calculated by summing all values in its corresponding row in the [ARROWS] matrix. This final score represents its overall dominance, and hypotheses were ranked accordingly.

### 3. Complex Scrutiny: Strategy Ranking

This analysis aimed to rank the overall quality of each complete diagnostic strategy (lottery).

- **Creation of the Granularity Matrix ([GRANU])**:
  - **Objective:** To capture the ‘granularity’ of comparisons between lotteries by counting the number of hypotheses for which one lottery dominates another.
  - An *M x M* matrix (where *M* is the number of lotteries) was created. For each pair of lotteries *(i, l)*, the cell *GRANU(i,l)* was populated with the number of hypotheses for which lottery *i* had a strictly greater plausibility score than lottery *l*. This measures the ‘depth’ of dominance.
- **Creation of the Popularity Matrix ([POPU])**:
  - **Objective:** To simplify the ordering relationship into a binary (win/loss) outcome between lotteries, without regard to the granularity of the comparisons.
  - An *M x M* matrix was created from the [GRANU] matrix. The cell *POPU(i,l)* was set to 1 if *GRANU(i,l) > 0*, and 0 otherwise. This represents a simple binary ‘win/loss’ outcome for each lottery duel.
- **Calculation of Final Strategy Ranking**: The final Popularity Score for each lottery was the sum of its row in the [POPU] matrix. The final Granularity Score was the sum of its row in the [GRANU] matrix. These scores were then used to calculate the average strategic performance of each of the four experimental groups.

### 4. Graphical Visualisation

- **Chord Diagram Preparation**: The results of the scrutinies are formatted into a standardised JavaScript matrix. Each object in the matrix represents an ordering relationship as a connection between a ‘source’ and a ‘target’ object (e.g., ‘hypotheses’ for the simple Condorcet scrutiny, or ‘lotteries’ for the complex scrutinies), with an associated ‘value’.
- In each Chord diagram, an arrow with a thickness corresponding to its ‘value’ is drawn from the ‘source’ to the ‘target’.
- The matrix is then inserted into an HTML template file to generate a Chord diagram for each subgroup.

